# Hao-Fountain Syndrome: 32 novel patients reveal new insights into the clinical spectrum

**DOI:** 10.1101/2023.11.14.23298551

**Authors:** Moritz Claudius Wimmer, Heiko Brennenstuhl, Steffen Hirsch, Laura Dötsch, Samy Unser, Pilar Caro, Christian Patrick Schaaf

## Abstract

Hao-Fountain syndrome (HAFOUS, OMIM: #616863) is a neurodevelopmental disorder caused by pathogenic variants in the gene *USP7* coding for USP7, a protein involved in several crucial cellular homeostatic mechanisms and the recently described MUST complex. The phenotype of HAFOUS is insufficiently understood, yet there is a great need to better understand the spectrum of disease, genotype-phenotype correlations, and disease trajectories.

We now present a larger cohort of 32 additional individuals and provide further clinical information about six previously reported individuals. A questionnaire-based study was performed to characterize the phenotype of Hao-Fountain syndrome more clearly, to highlight new traits, and to better distinguish the disease from related neurodevelopmental disorders. In addition to confirming previously described features, we report hyperphagia and increased body weight in a subset of individuals. HAFOUS patients present an increased rate of birth complications, congenital anomalies, and abnormal pain thresholds. Speech impairment emerges as a potential hallmark of Hao-Fountain syndrome. Cognitive testing reports reveal borderline intellectual functioning on average, although some individuals score in the range of intellectual disability.

Finally, we created a syndrome-specific severity score. This score neither indicates a sex-nor age-specific difference of clinical severity, yet highlights a more severe outcome when amino acid changes colocalize to the catalytic domain of the USP7 protein.

## Introduction

Ubiquitin-specific protease 7 (USP7), also known as Herpesvirus-associated protease (HAUSP), plays a crucial role as a regulator of ubiquitination in genome stability pathways ^1^. Recently, it was also described to contribute to the MUST pathway, which regulates intracellular protein trafficking and recycling ^2,3^. Another protein involved in this pathway, MAGEL2, is associated with Schaaf-Yang (SYS, OMIM #615547) ^4^ and Prader-Willi syndromes (PWS, OMIM #176270)^5^, and USP7 haploinsufficiency was subsequently shown to also cause a neurodevelopmental disorder ^3,6^. The initial report included seven individuals, six cases with genomic deletions containing the *USP7* gene and one individual with a *USP7* nonsense mutation^3^. Patients presented with developmental delay (DD), autism spectrum disorder (ASD), seizures, a behavioral phenotype, and signs of hypogonadism. Intriguingly, this clinical presentation shows great overlap with PWS and SYS by neurodevelopmental delay, failure to thrive, endocrine disruptions, behavioral anomalies, and muscular hypotonia^5,7–9^.

To date, the largest cohort of patients affected by Hao-Fountain Syndrome (HAFOUS, OMIM: #616863) was presented by Fountain et al. in 2019 describing 16 novel patients ^6^. The consistency of symptoms, regardless of the underlying variant type, supported the concept of USP7 haploinsufficiency as the causal factor of the disorder. Individuals presented with DD/ID, eye anomalies, feeding difficulties, GERD, behavioral anomalies, and ASD as the most frequent features. Furthermore, an in-depth secondary analysis of brain MRI images revealed various morphological anomalies with white matter paucity as the most frequent finding, reflected by a significant reduction of thickness of both splenium and body of the corpus callosum^6^.

In subsequent case reports, six more patients were added to the publicly available cohort ^10–13^. These new cases implicate a higher variability of the clinical spectrum, as fetal complications^10,11^, congenital heart defects ^10^, and isolated tubal torsion ^12^ were first described. Next to these specific items, the clinical presentation of these patients is mainly according to the phenotype described previously.

We here present 32 novel individuals with variants in *USP7* and provide further clinical information about six previously described patients with a first report on intellectual functioning, growth parameters, and pregnancy complications. Finally, by creating a severity score of clinical phenotype, we provide an outline of genotype-phenotype correlations.

## Subjects and Methods

This study is based on a 12-page questionnaire sent to all families listed in the USP7 registry. This registry was built in cooperation with the Foundation for USP7-related Diseases (www.USP7.org), enrolment was based on genotype. We included all individuals with confirmed genetic variants affecting the *USP7* gene and categorized patients based on type of mutations. Variants, including cases from the literature, were standardized using Mutalzyer 3 ( https://mutalyzer.nl) and re-classified according to the AMP/ACMG criteria ^14^. The questionnaire used within this study was designed with the Foundation for USP7-related Diseases to capture previously unrecognized symptoms. It was conceived to be filled out by the parents and always contained the option to select "unknown" if reporting was impossible. Growth centiles and z-scores were calculated using an online tool based on CDC standard values ( https://www.pedz.de/en). Detailed clinical reports were requested, especially brain imaging and EEG reports. The data presented in this study are primarily based on parental reports. We endeavored to enhance the validation process by incorporating clinical reports whenever feasible. It should be noted that results from testing reports, radiological imaging reports, and behavioral assessments were accepted without secondary evaluation. Thirty-eight families replied to the request and sent back at least the questionnaire and a genetic testing report, accompanied by a consent form. Six of those patients had been previously reported by Fountain et al. in 2019 ^6^. We labeled the cases based on their chronological appearance in the literature and therefore report individuals #30 to #61 (*Supplemental table 1*).

If not stated otherwise, the frequency of the symptoms always describes the prevalence in all publicly accessible cases, including patients described by Hao et al., Fountain et al., Capra et al., Zampieri et al., Zheng et al., and the participants of this study^3,6,10,12,13^.

A severity score was developed based on the most frequently occurring clinical features. Where available, the score considers the range of severity, e.g., one or two points based on the severity of the developmental delay. A scale of 0 to 24 points was assigned and normalized afterwards to avoid bias by data availability. The exact procedure next to the calculation of individual severity scores can be seen in the supplemental material (*supplemental methods, supplemental table 14* ). Severity scores were calculated exclusively for participants of our study. The categorization of patients with a missense variant, as described below, was conducted based on the protein domain boundaries outlined by Valles et al. in 2020^1^.

Differences between two groups for the severity index were evaluated using Student’s t test. Whenever comparing more than one group for continuous variables (developmental milestones, growth parameters), a one-way ANOVA analysis was performed. One-sample t tests were performed in a z-normalized setting to assess deviation from the average population. Analyses were conducted using GraphPad (Version 9.5.1) or R (R Core Team, Vienna, Austria), and data for all patients were collected in IBM SPSS Statistics 29.

## Results

We present 32 novel patients (16 M/16 W) with an average age of 9.97 years (2-20 years). All individuals harbor variants affecting *USP7* (two whole-gene deletions, three partial gene deletions, one intragenic deletion, six truncating variants, 15 missense variants, and five splice-site variants) ( *Table 1*). Figure 1 illustrates the distribution of missense and nonsense variants, revealing an enrichment of missense variants within the catalytic domain. Notably, no N-terminal nonsense variants were reported. The symptom reported most frequently is motor developmental delay, reflected by an overall delay in attainment of developmental milestones, followed by motor impairment, gait abnormalities, and muscular hypotonia. Furthermore, autism spectrum disorder (ASD), communication deficits, an abnormal pain threshold, gastrointestinal issues, and brain MRI anomalies are typical features presented by more than two-thirds of the cohort analyzed in this study. A summary of the most frequent symptoms is provided in *Table 2*, and a deeper analysis of all assessed symptoms subdivided by genotype can be found in the supplemental material (*Supplemental Tables 1-13*).

**Figure 1:**
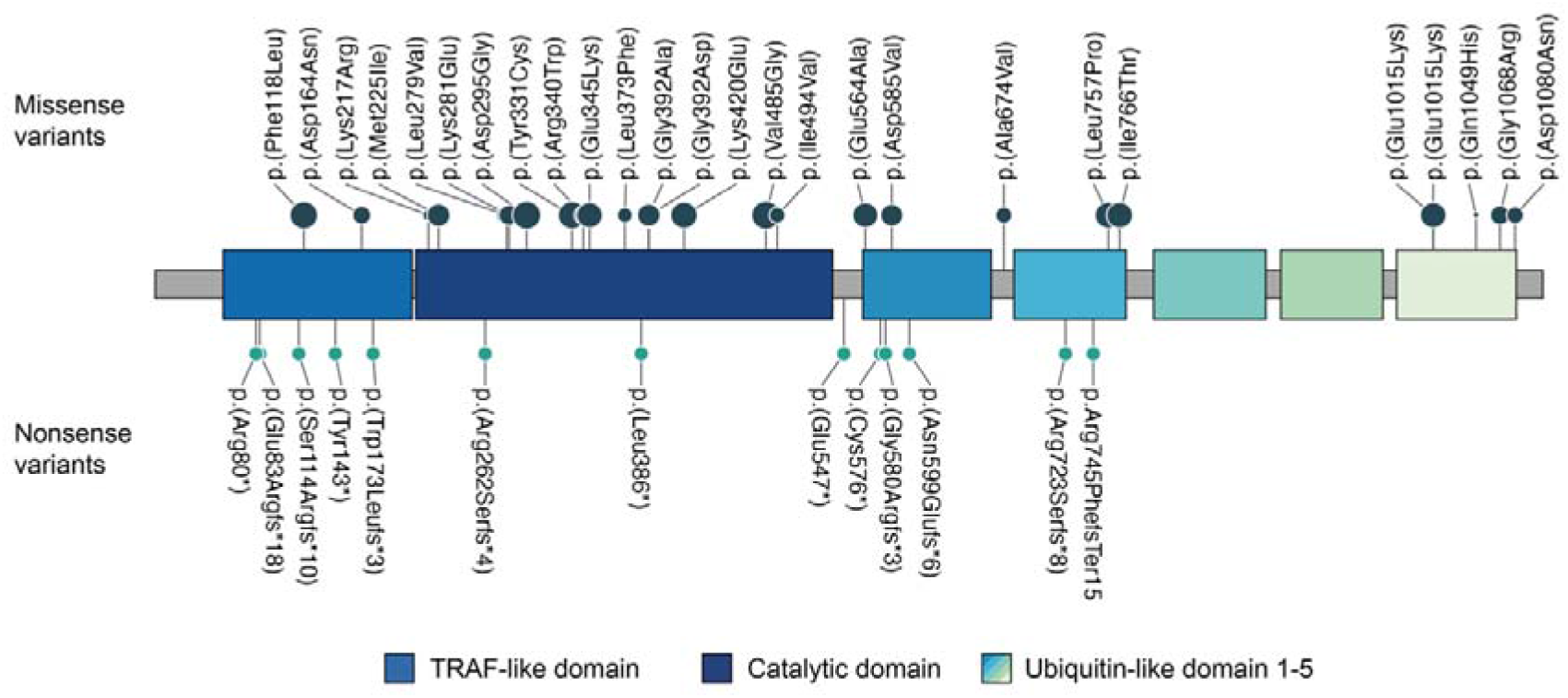
Distribution of *USP7* missense and nonsense variants. The known domains and the distribution of missense/nonsense variants along a two-dimensional representation of the USP7 protein are shown. The size of the points of the missense variants correlates with the respective REVEL score.

**Table 1.**
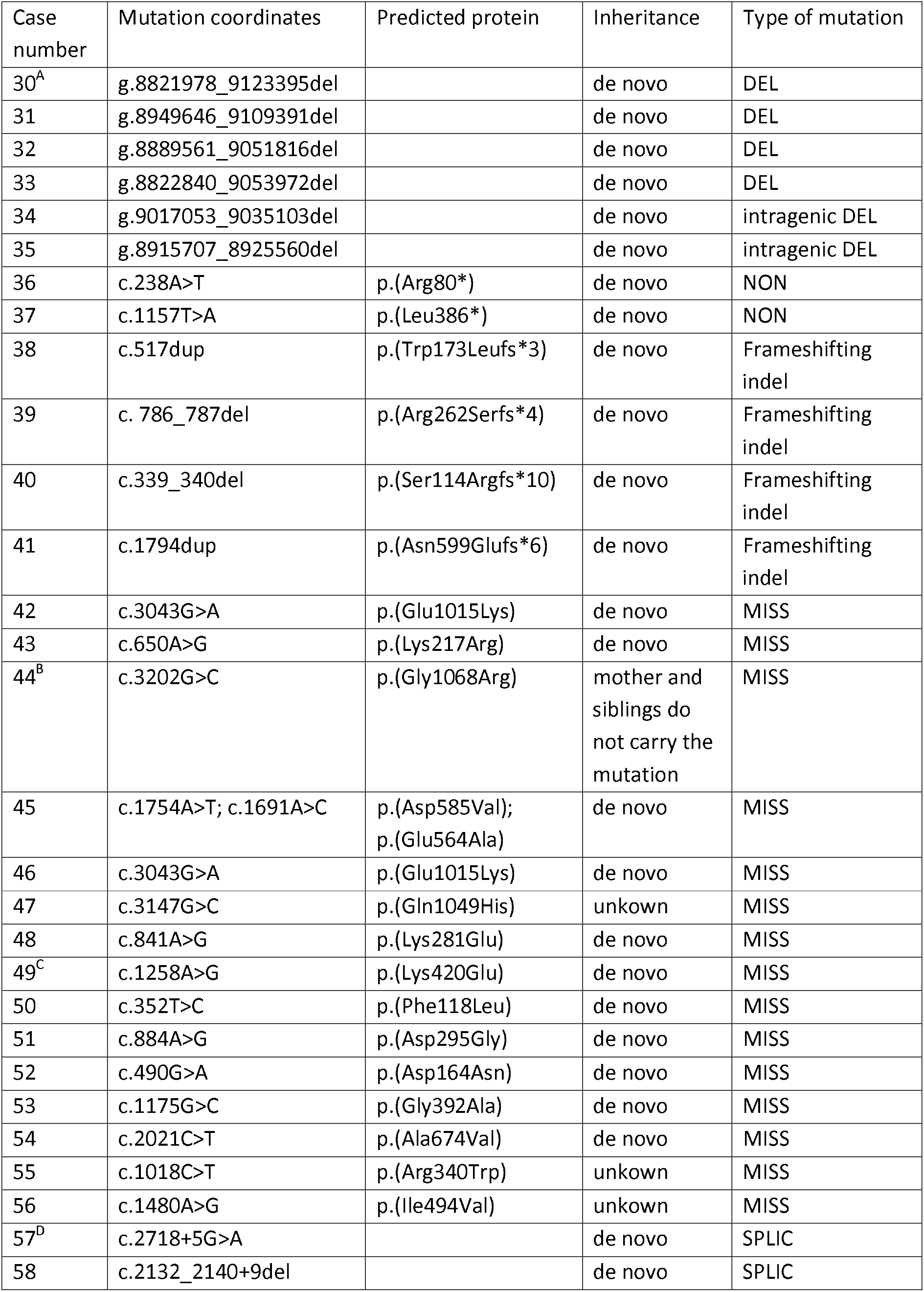
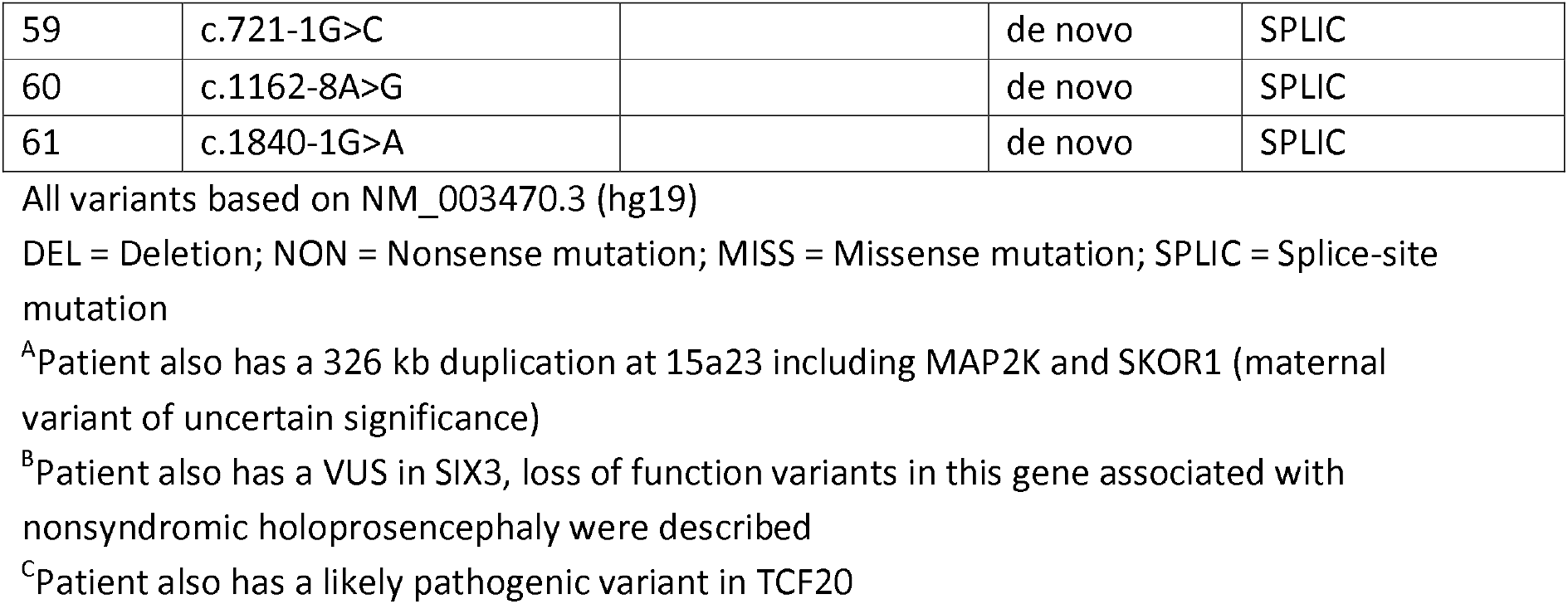
Novel cases with pathogenic USP7 variants.

**Table 2.** Summary of published and novel cases with pathogenic USP7 variants.

| **Clinical feature** | **Literature** | **Our study** | **Summary** |  |
| --- | --- | --- | --- | --- |
| Muscular hypotonia | 16/23 | 26/30 | 42/53 | **79 %** |
| Oral motor skills delayed/ impaired | 6/7 | 23/31 | 29/38 | **76 %** |
| Altered threshold for pain | 6/6 | 20/28 | 26/34 | **76 %** |
| Gross motor skills delayed/ impaired | 6/7 | 23/32 | 29/39 | **74 %** |
| ASD | 14/18 | 17/25 | 31/43 | **72 %** |
| Fine motor skills delayed/ impaired | 5/6 | 20/31 | 25/37 | **68 %** |
| Brain MRI anomalies | 26/21 | 7/13 | 23/34 | **68 %** |
| Abnormal gait | 8/17 | 23/30 | 31/47 | **66 %** |
| Limited communication skills | 2/6 | 22/31 | 24/37 | **65 %** |
| Neonatal hypotonia | 10/20 | 15/24 | 25/44 | **57 %** |
| Dental issues | 4/6 | 16/30 | 20/36 | **56 %** |
| Feeding difficulties | 14/22 | 16/32 | 30/54 | **56 %** |
| Chronic constipation/ diarrhea | 14/20 | 15/32 | 29/52 | **56 %** |
| Eye anomalies | 17/25 | 11/28 | 28/53 | **53 %** |
| Complications during pregnancy | 3/8 | 17/30 | 20/38 | **53 %** |
| ADHD | 7/17 | 14/25 | 21/42 | **50 %** |
| Sleep disturbances | 7/18 | 15/29 | 22/47 | **47 %** |
| Eating difficulties | 4/7 | 12/29 | 12/36 | **44 %** |
| Temperature instabilities | 2/4 | 12/30 | 14/34 | **41 %** |
| Small feet | 7/21 | 12/25 | 19/46 | **41 %** |
| Complications during delivery | 2/6 | 13/31 | 15/37 | **41 %** |
| Congenital anomalies | 2/5 | 12/29 | 14/34 | **41 %** |
| Seizures | 13/27 | 10/30 | 23/57 | **40 %** |
| Neonatal poor suck | 6/19 | 14/31 | 20/50 | **40 %** |
| Hyperphagia | 1/5 | 13/31 | 14/36 | **39 %** |
| Incontinence | 3/6 | 12/32 | 15/38 | **39 %** |
| GERD | 10/19 | 8/30 | 18/49 | **37 %** |
| Behavioral problems | 2/5 | 8/24 | 10/29 | **34 %** |
| Small hands | 8/21 | 6/22 | 14/43 | **33 %** |
| Scoliosis/ Kyphosis | 7/20 | 7/27 | 14/47 | **30 %** |
| Sleep apnea | 2/6 | 8/28 | 10/34 | **29 %** |
| Gaining weight difficulties | 7/19 | 8/32 | 15/51 | **29 %** |
| Contractures | 7/19 | 4/26 | 11/45 | **24 %** |
| Hypogonadism | 7/19 | 4/28 | 11/47 | **23 %** |
| SGA | 4/7 | 4/29 | 8/36 | **22 %** |
| EEG anomalies | 1/5 | 3/15 | 4/20 | **20 %** |
| Skin picking | 4/22 | 5/30 | 9/52 | **17 %** |
| LGA | 0/7 | 6/29 | 6/36 | **17 %** |
| Short stature | 4/19 | 2/29 | 6/48 | **13 %** |
| GH deficiency | 2/4 | 0/20 | 2/24 | **8 %** |

### Growth parameters

Growth parameters from 39 patients indicate a trend towards increased height, weight, and BMI, with BMI achieving a statistically significant level (one sample t test; p _BMI_= 0,0377). Among the entire cohort, the mean weight measures at the 59 ^th^ centile, the mean body height measures at the 60 ^th^ centile, and the mean BMI (18.84) lies within the 61 ^st^ centile (*Figure 1, Table 3*). For 34 total cases, we could analyze an age-adjusted BMI percentile. One case is below the 5 ^th^ centile indicating underweight as defined by the CDC ^15^, 22 cases are in the average weight range, six fall in the overweight range, and five fulfill obesity criteria. Therefore, 32 % of our entire cohort are overweight or obese. Although there is no general trend towards growth retardation, 13 % (6/48) of this patient cohort indicate short stature, defined as a body size at least two standard deviations below an age-adjusted standard population^16^. Furthermore, 14 of 43 (33 %) patients showed small hands, while 19 of 46 (41 %) showed small feet, in both cases defined as feet and hand length below the age-adjusted 25^th^ percentile (for novel cases).

**Table 3.** Developmental/ speech milestones, growth parameters, and IQ of patients with Hao-Fountain Syndrome.

|  | **Mean value** | **Expected value**<br>**(normal population)** | **Assessed individuals**<br>**(n)** |
| --- | --- | --- | --- |
| First spoken word | **21,1** months | 18 months | *30* |
| First combining words | **32,9** months | 24 months | *24* |
| Sit w/o support | **11,7** months | 6 months | *46* |
| Crawl | **14,3** months | 12 months | *27* |
| Walk w/o support | **26,6** months | 15 months | *47* |
| FSIQ | **78,30** | 100 | *10* |
| Body weight percentile | **58,35** | 50 | *39* |
| Body length percentile | **59,23** | 50 | *38* |
| BMI | **18,84** | depends on age | *36* |
| BMI percentile | **60,63** | 50 | *34* |
| Birth weight percentile | **41,19** | 50 | *36* |
| Birth length percentile | **46,67** | 50 | *38* |

### Pregnancy and birth

Contrary to the body measurements described above, patients demonstrated a trend towards reduced birth weight and body size. Average birth weight is at the 42 ^nd^ centile, and mean birth size at the 47 ^th^ (*Figure 1, Table 3).* Neither sex nor genotype appear to influence growth parameters or birth body measurements ( *Supplemental Figures 1-3* ). Interestingly, we see both a subgroup being small for gestational age (birth weight below the 10 ^th^ centile) and one being large for gestational age (birth weight above the 90 ^th^ centile). The former criterion is met in 8 of 37 cases (22 %), the latter in six of 37 cases (16 %).

Neonatal muscular hypotonia affected a majority of 25 out of 44 individuals (57 %). Head circumference measures at the gestational age-adjusted 47 ^th^ centile and does not indicate altered head circumference at birth. A single patient (3%) manifested hydrocephalus as an infant. Twenty of 38 parents surveyed (53%) reported problems during pregnancy, which ranged from common pregnancy-related issues such as gestational diabetes to detected cardiac peculiarities or polyhydramnios. Decreased fetal movement was reported in seven of 44 cases (16 %). While preterm delivery occurred in eight of 40 cases (20 %), the mean gestational age was 37.77 weeks. 15 of 37 parents (41 %) reported complications after delivery and described those in a free-text answer, at which temperature instability was named most often (five times), followed by severe jaundice (four times) and respiratory distress (three times). Described congenital malformations met the criteria of having significant medical, social, or cosmetic consequences for the affected individual in 14 of 34 cases (41 %). After the first case of a HAFOUS patient with congenital anomalies was described by Capra et al. in 2019, we can now add 12 more individuals presenting anomalies of the cardiac system (four times), the motor system (3 times), dysmorphisms of the head (3 times), malformation of the oral cavity or pharynx (3 times), and one patient born with a duplex pelvicalyceal system in the right kidney.

Given the disparities between current body measurements and those taken at birth, our aim was to explore the potential positive correlation between age and body weight, height, and BMI. Surprisingly, we observed a noteworthy decline in body weight and BMI percentiles as age increased (Supplemental Figure 4, A-C). Nevertheless, in our cohort and in line with the general population, absolute BMI values exhibit a positive correlation with age (Supplemental Figure 4D).

### Developmental milestones

According to WHO data, the first motor development milestone, "sitting without support," is reached after 6.0 months, while "hand-and-knees crawling" is reached after 8.5 months, and "walking alone" can be accomplished after 12.1 months of life in the general population ^17^. Presenting a delay at every motor development milestone, the cohort described here reached free sitting on average after 11.7 months, crawling after 14.3 months, and free walking after 26.6 months ( *Table 3*). Twenty-six of 46 patients reached unsupported sitting after more than 9.2 months, 14/27 patients reached crawling after more than 13.5 months, and 39/47 patients reached free walking after more than 17.6 months. Each of these mentioned time points represents the respective 99^th^ centile in the general population.

Regarding speech developmental milestones, children are expected to speak their first word at 18 months and can build the first two-words sentences 24 months after birth ^18^. The cohort described here spoke their first words on average after 21.1 months and built their first two-word sentence after 32.9 months ( *Table 3*). In this study, the oldest patient being nonverbal was 14 years old and can also not safely walk without support (#43), while Fountain et al. have already described a 20-year-old patient as nonverbal.

### Intellectual ability

Fifteen families provided intellectual testing reports, which featured a full-scale intelligence quotient in 10 cases and a description of severe ID in two cases (WISC or WPPSI testing procedures). The mean FSIQ for this cohort is 78.3 ( *Table 3* ) and falls within the range of borderline intellectual functioning. Nonetheless, two of 12 patients present an IQ below 35, which defines severe intellectual disability (ID). Four of 12 patients score between 50 and 69, indicating mild ID, two of 12 patients score between 70 and 85, indicating borderline intellectual functioning, and another four patients show normal intellectual functioning with an IQ above 85. The highest IQ measured in this cohort is 105. Two previously described patients scored in the range of moderate ID ^11,12^.

According to previously published data, an extensive chart and questionnaire review revealed signs of intellectual disability or (a history of) developmental delay in all cases.

### Behavioral characteristics

Behavioral characteristics were stated to be a defining feature of HAFOUS and, therefore, also queried meticulously in this study. Testing for autism spectrum disorder was conducted in 15 cases, leading to a diagnosis in 10 cases (67 %). To overcome a selection bias, parents were also asked to state whether their child showed autistic features. Thirty-one parents reported positive testing or autistic behavioral features (72 %), while 12 neither tested positive nor showed behavioral features indicative of autism. ASD is more prevalent than initially expected in HAFOUS. Five of 21 parents reported an improvement of the autistic features over time, eight of 21 saw a worsening, and eight of 21 could not observe any change.

Nine of 42 reported an attention deficit hyperactivity disorder (ADHD) diagnosis (21 %), another 12 of 42 described ADHD-like behavioral traits, but lack of a formal diagnosis (29 %), and 21 of 42 did not observe any feature reminiscent of ADHD.

Previous publications indicated manifold behavioral traits, including aggressive behavior, temper tantrums, impulsivity, compulsivity, stubbornness, and manipulative behavior. Improving the objectivity of this assessment, we collected answers on a *Likert scale* for different behavioral traits, which would typically not be expected to appear often. Defined between 1 indicating "always" and five indicating "never", the scores ranged from 4.18 ("destructive in her/his room") to 2.46 ("stubbornness"). Next to the latter, "impulsivity", "overly social", and "physical contact seeking" scored values closer to always than never. Previously described features like "aggressiveness" or "manipulative behavior" scored 3.46 and 3.95, respectively ( *supplemental figure 5* ). Therefore, we can only partially confirm the previously described behavioral anomalies.

Furthermore, 10 of 29 parents reported general behavioral problems (34 %), 16 of 31 patients showed obsessive-compulsive behavior (52 %), and only 9 of 52 parents observed skin picking (17 %).

### Musculoskeletal phenotype

One of the most frequent symptoms of HAFOUS is hypotonia, prevalent in 42 of 53 patients (79 %). Hypotonia improved over time in the vast majority of 23 out of 30 cases (worsened in two cases, no change in 5 cases). This is surprising, given that hypotonia at the time of examination is more prevalent than neonatal hypotonia, indicating a numerical increase of cases, although the individual courses may be ameliorating on average. Aside from low muscle tone, other dysfunctions of the muscular system can be observed. Abnormal gait was reported in 31 of 47 individuals (66 %), and 23 of 35 exhibit balance unsteadiness (66 %). Motor dysfunction affects all categories and is indicated by fine motor delay/impairment (25/37, 68 %), gross motor delay/impairment (29/39, 74 %), as well as oral motor skills delay/impairment (29/39, 76 %).

Less prevalent are scoliosis/kyphosis, which can be seen in 14 of 47 patients (30 %), and joint contractures, reported by 11 of 45 affected families (24 %), most frequently located at the lower extremity (6 cases).

Diverse dental issues were described by 20 of 36 parents (56 %), and the severity of the issues ranges from severe malformations to cosmetic conspicuities. "Large front teeth" were reported five times, while "deep palatal vault" and "supernumerary teeth" were reported three times, respectively. Seventeen more issues were described no more than one time.

### Neurological/ ophthalmological features

Eye anomalies were previously reported to affect a vast majority of HAFOUS patients. We can confirm this for 28 of 53 patients in this cohort (53 %) and found farsightedness (10/34, 27 %) to appear most frequently (*supplemental table 5*).

One of the queried items proposed by the foundation was an abnormal pain threshold, a known symptom of PWS^19^. We saw a high threshold for pain at 19 of 34 (56 %) patients and a low threshold for pain at another seven of 34 (21%) patients, indicating 76 % with a parent-reported dysfunctional pain threshold.

Although delayed speech development was described for HAFOUS previously and is confirmed here, speech abilities at a later stage have never been described. 24 of 37 (65 %) parents reported limited communication skills, being reflected both by reduced expressive (26/38, 68 %) and receptive (23/36, 64 %) communication. Furthermore, parents were asked which feature would be the most disabling symptom of their children and in 16 of all 38 questionnaires, parents reported speech problems as the most disabling feature, highlighting the significance of this deficit for the affected individuals (cognitive impairment named nine times, behavioral anomalies named eight times).

Hearing difficulties were reported in nine of 51 (18%) cases, and temperature instability was reported in 14 of 34 (41 %) patients.

Fountain et al. extensively analyzed brain MRI reports in 2019, and we can mainly confirm their findings based on the 13 new reports received. Combined with previously published reports, brain MRIs exhibit anomalies of the ventricular system in 12 of 25 cases, signs of white matter paucity/ small corpus callosum in 11/25 cases, simplified gyral patterns in 4/25 cases, white matter hyperintensities in 3/25 cases, vermian atrophy in 3/25 cases, and hippocampal anomaly as well as a thick corpus callosum one time respectively. Summarizing the reports, 23 of 34 (68 %) patients show any sign of abnormality described in their brain MRI report with a wide variety of clinical significance as described above.

### Seizures and sleep

Sleep disturbances were reported for 22 of 47 (47 %) patients and could partially explain the frequent occurrence of fatigue (26/37, 70%) and a great need for rest (23/36, 64%). Sleep apnea occurs in 10 of 34 (29 %) individuals.

Seizures were previously stated as a very prevalent and characteristic feature of HAFOUS. 23 of 57 (40 %) patients in the cohort described here were affected by seizures or have a history of seizures; most effectively treated with anti-epileptic medications. The highest frequency of seizures reported is 14 per year. Three of 32 (9 %) individuals had a febrile seizure. In most cases, the reason for the seizures could not be clarified. Three of 19 (16 %) EEG reports we received showed anomalies, in all three cases describing (probable) epileptic potential. Ten parents provided information about the course of this symptom, and while four did not observe any difference, six families confirmed an improvement concerning seizures over time.

### Gastrointestinal features

This cohort’s most frequent gastrointestinal issues are constipation or diarrhea, affecting 29 of 52 (56 %) patients. Incontinence inappropriate for the individual’s age was reported by 15 of 38 (39 %) parents. Gastroesophageal reflux disease can be observed in 18 of 49 (37 %) cases, and five of 35 (14 %) parents reported jaundice beyond the neonatal period. In four of 38 (10 %) cases, gastrointestinal issues were described as the most disabling symptom.

### Eating behavior

Abnormal eating behavior is one of the key features of PWS and, therefore, also of great interest in HAFOUS research. 30 out of 54 individuals (56 %) in the cohort here experienced feeding difficulties during infancy, closely associated with 15 of 51 (29 %) patients having difficulties gaining weight. Eating difficulties can be observed in 16 of 36 (44 %) cases. For the first time, we describe 14 of 36 (39 %) HAFOUS patients presenting hyperphagia. While this parental assessment lacks objectivity, further behavioral characteristics like mouthing behavior (10/34, 29 %) and chewing of inedible objects (10/37, 27 %) support this finding. Furthermore, 23 % (3/13) of the children showing hyperphagia are obese, while only 5 % (1/20) fulfill this criterion in the non-hyperphagic group. Manifestation of hyperphagia began three times in the neonatal period (first month of life), four times in infancy (second month until the end of the second year), five times in early childhood (three until eight years), and once later than that.

### Further physical characteristics

We also asked for endocrinological and respiratory dysfunction but could not identify any features of high prevalence.

Hypothyroidism (1/21, 5 %), adrenal insufficiency (1/21, 5 %), and growth (GH) deficiency (2/24, 8 %) were reported in only few patients, while hypogonadism was observed in 11 of 47 patients (23 %). We also included 4 of 16 boys with cryptorchidism/ micropenis.

Only a few patients depended on respiratory assistance in the form of intubation (2/35, 6 %) or mechanical ventilation (3/35, 9 %). In 2 of 3 cases, chart review revealed that ventilation was necessary immediately postnatally, while the context could not be outlined for one further case. Asthma was prevalent in 8 of 36 (22 %) patients.

Eczema was described by 12 of 34 (35 %) parents.

### Genotype-phenotype association

The cause of clinical heterogeneity in *USP7*-related disorder is not very well understood. Aiming to overcome this issue, a severity score was developed as described before (methods, supplemental methods). Anderson-Darling test (p=0.4699), Shapiro-Wilk test (p=0.3424), and Kolmogorov-Smirnov test (p>0.1) were passed. Therefore, normal distribution can be assumed ( *supplemental figure 6A* ). The mean severity score is 0.4937 (SD: 0.1561) for the 38 values we generated.

To better understand influencing factors on the severity, we compared both sexes and found no difference (M: 0.49 SD 0.14, n:16; W: 0.50 SD 0.17, n:22) ( *figure 2A).* Furthermore, age and type of the underlying genotype do not influence severity significantly (DEL: 0.47 SD 0.13, NON: 0.41 SD 0.12, MISS: 0.53 SD 0.18, SPLIC: 0.49 SD 0.07) ( *figure 2B, supplemental figure 6B* ). Due to the heterogeneity of severity within the group harboring missense variants, a correlation of exact position of the mutation and severity was investigated. Missense mutations were grouped based on functionally relevant protein regions. A mean value of 0.64 (n: 10) was found for the catalytic domain, 0.48 (n: 2) for the TRAF-like domain, and 0.41 (n: 8) for the UBL-like domain, reflecting a significantly increased severity score for mutations affecting the catalytic domain of USP7 (figure 2C, p = 0.0041).

**Figure 2:**
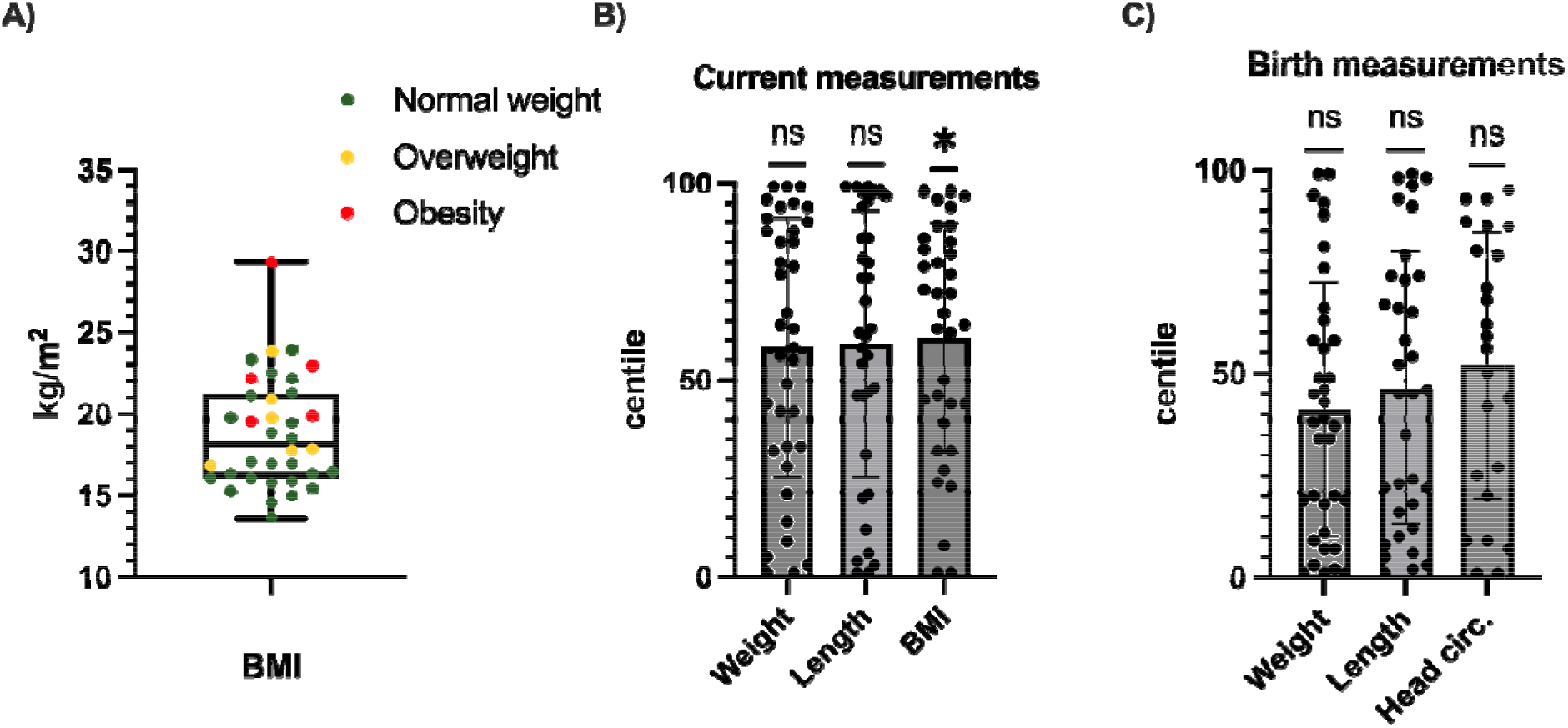
Body measurements of individuals with Hao-Fountain Syndrome; one sample t-test performed for a mean value of 50. A: BMI subdivided by normal weight, overweight, obesity; B: Current measurement centiles; C: Birth measurement centiles

**Figure 3:**
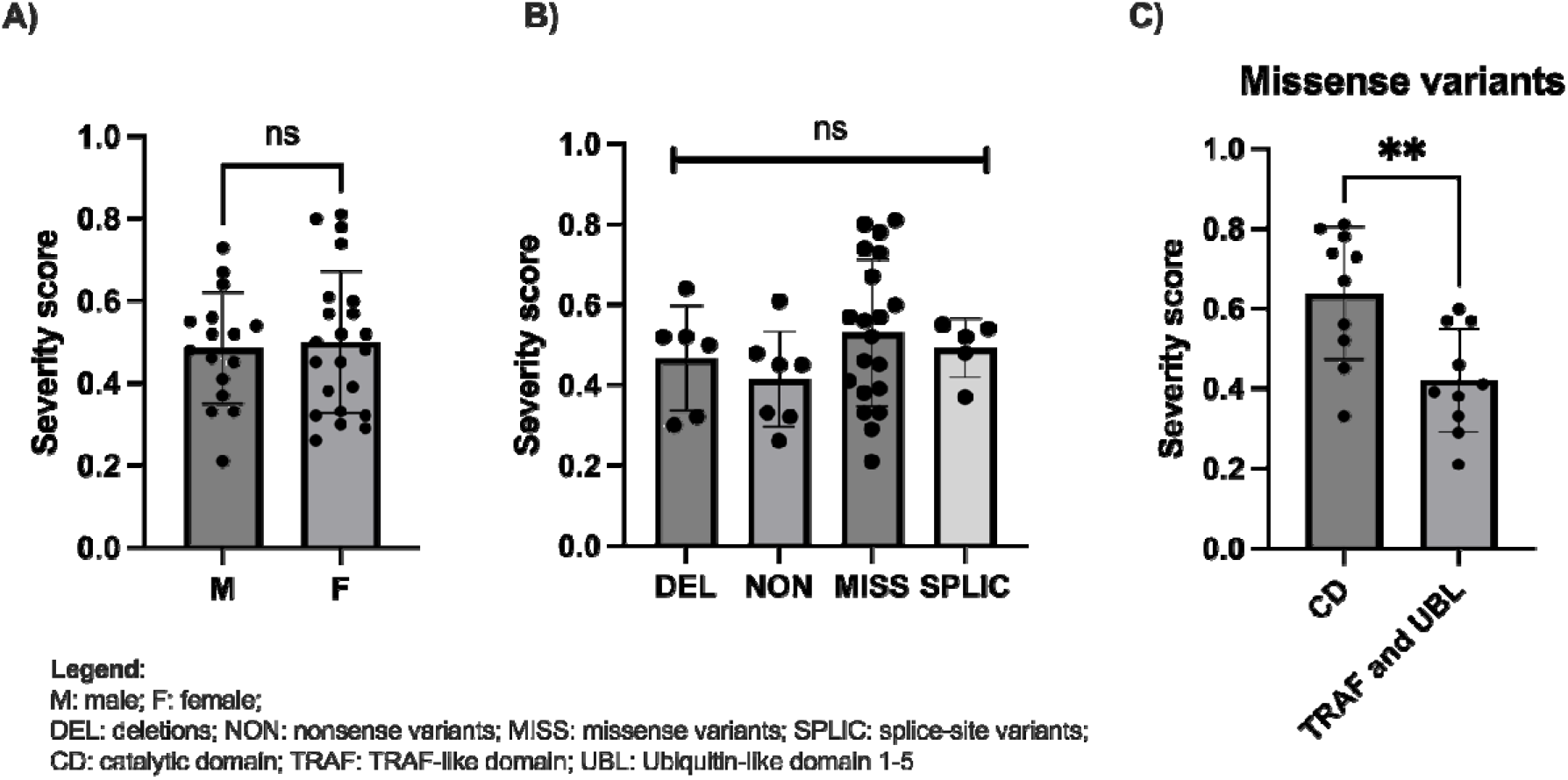
Assessment of severity score values in variable subgroups; Student’s t-test and one-way ANOVA. A: Severity score plotted for sex; B: Severity score for each type of mutation.; C: Severity score of missense mutations grouped by localization (Catalytic domain vs. TRAF-like- and UBL-domain).

Furthermore, to assess the predicted effect of missense variants, commonly used prediction scores (CADD v1.6, M-CAP v.3.5a, REVEL v.3.5a21-23) were annotated for all biologically possible USP7 missense variants and mapped onto a linearized representation of the USP7 protein, as previously demonstrated ^20^. A generalized additive model was built using the geom_smooth function of the R (R Core Team, Vienna, Austria) ggplot2 package to plot a smoothened line and confidence interval (Supplemental Figure 7). Violin plots of pathogenicity scores associated with all biologically possible missense variants falling into either the catalytic domain, the TRAF-like domain or one of five Ubiquitin-like domains were generated. Missense variants in catalytic domain are associated with statistically significant higher pathogenicity scores shown by the assessment of three individual scores (REVEL, MCAP, and CADD score) (Supplemental Figure 8).

## Discussion

HAFOUS is a neurodevelopmental disorder leading to a variable clinical spectrum in terms of occurrence and severity of symptoms. Speech and motor delay are most prevalent. Patients typically present with communication deficits, muscular hypotonia, motor issues, abnormal threshold for pain, and autism spectrum disorder. Complications during birth and congenital anomalies are frequently present. Further symptoms such as seizures, hyperphagia, or scoliosis only affect a subset of individuals. As previously proposed, this disorder can be considered part of a disease family with SYS and PWS ^21^, showing a wide overlap of symptoms between these disorders and a hypothesized shared path of pathophysiology via the MUST complex ^22^. Developmental delay, muscular hypotonia, reduced birth weight, and infant feeding difficulties are shared features among these disorders^5,7^.

Furthermore, endocrine dysfunction, also described as hypothalamic insufficiency, is a crucial feature of SYS and PWS, but could not be seen frequently in our cohort ^23,24^. ASD is much more prevalent in HAFOUS and SYS (72 % and 78 %) compared to PWS (27 %) ^7,25^. SYS, with its unique prevalence of joint contractures and respiratory dysfunctions accompanied by the most severe manifestation of developmental delay, intellectual disability, and increased mortality, can be considered the most severe disorder within this disease family ^7^. The causative factors need to be further investigated, but a neomorphic function of the truncated MAGEL2 protein or a suppression of leaky expression from the maternal allele at SYS was previously hypothesized to contribute to the alteration of manifestation between these disorders ^22^. HAFOUS positions itself as the least severe manifestation within this disease family due to milder developmental delay, higher intellectual functioning, and mostly an absence of endocrine dysfunctions (see ^7,26,27^). Considering the MUST pathway as one of the driving factors of these diseases, it is noteworthy to mention that pathogenic variants in *USP7* were reported only heterozygous, suggesting the presence of wild-type and variant USP7 proteins, while MAGEL2 mutations associated with PWS and SYS are functionally hemizygous, suggesting the complete absence of wild-type MAGEL2. Therefore, it can be presumed that mutations underlying SYS or PWS have a greater impact on the functionality of the MUST complex than mutations causing HAFOUS. Nevertheless, the variability of symptoms is intriguing, and some patients face a much more severe phenotype than others. Although we provide data indicating the most severe phenotypes in patients with missense variants within the catalytic domain, predicting the severity of phenotypes based on individual mutations remains challenging.

This work sheds light on new, previously undescribed features of HAFOUS. Most remarkably, we found an abnormal threshold for pain affecting 76 % of the cohort. Furthermore, we describe some individuals presenting hyperphagia and potentially associated obesity. Complications during pregnancy and congenital anomalies of varying severity had not been reported previously. In addition to describing symptoms for the first time, we provide more profound data for all previously described symptoms and observe a higher prevalence of ASD than indicated by previous studies. Some features like seizures and particularly aggressive and impulsive behavior are not as prevalent as initially hypothesized.

Developmental anomalies manifest in a variable trajectory of body weight, height, and BMI. In contrast to the general population, we note slightly diminished birth height and weight, with current body measurements reflecting a modest increase. However, there is a discernible decrease in BMI and body weight with advancing age, suggesting an irregular and atypical pattern of body growth.

Most significantly, we also analyzed genotype-phenotype correlations based on a developed severity score and found increased severity scores for patients with a missense mutation inside the catalytic domain of *USP7*, compared to those with a mutation outside the catalytic domain. Evaluation of in silico scores revealed significant higher pathogenicity scores if missense mutations occurred in the catalytic domain. Based on these results, we hypothesize that missense variants within the catalytic domain of the protein are associated with a more severe clinical phenotype compared to variants in the other parts of the protein. We recommend analyzing larger cohorts even in the initial phase of investigating a novel disease to ensure a reliable foundation of data. Our approach to include the disease community when creating the questionnaire, appears helpful for gathering data on previously undescribed features.

TRIM27 is another component of the MUST-complex and was not previously described in case reports or systematic patient cohort analyses of neurodevelopmental disorders. This is surprising as other proteins of this pathway can be assigned to various neurodevelopmental conditions^22^. We currently aim to identify individuals carrying *TRIM27* variants to provide a systematic analysis.

Interestingly, a recent paper implicates all the variable functions of USP7, but especially contribution of USP7 to epigenetic transcriptional regulation, in the search for an etiology of HAFOUS^28^. This seems reasonable to us as this group revealed that USP7 is highly associated with histone ubiquitination on a transcriptional and posttranslational level as a silencer or stabilizing partner for relevant factors of the PRC1 complex and therefore contributes to the expression of several neurodevelopmental factors like AUTS2 ^28^. Establishing animal models of this disorder could help understand the exact mechanism of pathophysiology and the cellular pathways contributing to the phenotype. Remarkably, an overexpression mouse model of *Usp7* presented altered dendritic arborization, neuronal migration, a behavioral phenotype, and altered pain threshold, which is partially reminiscent of the phenotype observed in our cohort suggesting dosage-sensitivity of Usp7 during neurodevelopment^29^.

This study is subject to some limitations. The symptoms were recorded in a questionnaire by interviewing the parents and are thus primarily subjective. Whenever possible, clinical charts and records were assessed to ensure objectivity. Furthermore, the developed severity score is limited by a detection bias, meaning that the objective nature of such scores always limits the possible coverage of symptoms and individuality.

A phenotyping study of individuals at the University Hospital of Heidelberg is currently being conducted to overcome this issue. By analyzing 32 novel and six previously described patients, this study helps understand the clinical spectrum of HAFOUS, reveals undescribed symptoms, and provides a first approach to a genotype-phenotype correlation. With this, we intend to contribute to a more reliable and sound genetic consulting and prediction of phenotype for affected individuals and families.

## Supporting information

Questionnaire

## Ethical statement

This study was performed under the research protocol H-34578, approved by the Institutional Review Board for Baylor College of Medicine and Affiliated Hospitals. All individuals and their guardians were informed and provided consent for the study.

## Statement of contribution

Methodology: MCW, HB; Investigation: MCW, LD, PC; Validation: SH, SU; Project administration: MCW, PC; Writing – original draft: MCW; Writing – review and editing: CPS, MCW; Visualization: HB, MCW; Supervising: CPS

## Acknowledgments

The authors thank the individuals and the families with Hao-Fountain syndrome, who have provided clinical data and overwhelming support for our efforts. The authors also thank Ferdinand Althammer and Jannis Bücking for comments that greatly improved the manuscript.

## Conflicts of interest

The authors declare no competing interests.

## Data Availability statement

The authors confirm that the data supporting the findings of this study are available within the article and its supplementary materials.

## 7 Legends

**Supplemental figure 1:** Body measurements compared by sex

A: BMI values (Student’s t-test).; B: Current body measurement percentiles; C: Birth body measurement percentiles

**Supplemental figure 2:** Current body measurements compared by genotype

A: BMI values (One-way ANOVA); B: Current body length percentile (One-way ANOVA); C: Current body weight percentile (One-way ANOVA); D: BMI percentile (One-way ANOVA)

**Supplemental figure 3:** Birth body measurements compared by sex and genotype

A: Birth body weight percentiles (One-way ANOVA); B: Birth head circumference percentiles (One-way ANOVA); C: Birth body length percentiles (One-way ANOVA); D: Birth measurements compared by sex

**Supplemental figure 4:** Body measurements plotted against age indicating a significant impact of age on weight percentile (B), BMI percentile (C), and BMI values (C)

**Supplemental figure 5:** Behavioral traits of Hao-Fountain syndrome patients Several behavioral traits and parental-reported frequency of occurrence ranging from 5 (Always) to 1 (Never) (n: 36).

**Supplemental Figure 6:** Severity score QQ plot and assessment for different subgroups A: QQ plot proofing normal distribution of severity scores within our cohort; B: Severity score plotted against age of individuals (slope: 0,0004920) indicating no impact of age on severity; C: Severity score of patients with a truncating mutation plotted for the location of the predicted location of the truncation; D: Severity score of patients with a missense mutation plotted for the location of the predicted location of the truncation (Tukey’s multiple comparison test following one-way ANOVA; CD vs. UBL: adjusted p-value = 0.0131)

**Supplemental Figure 7:** Comparison of three pathogenicity score predictions

**Supplemental Figure 8:** Statistical comparison of domain-associated pathogenicity scores

**Supplemental table 1 – Molecular phenotype:** Comprehensive molecular phenotype information on known patients with HAFOUS, including variant coordinates, inheritance, and mutational group allocation, in silico scores, and AMP/ACMG re-classification (all based on NM_003470.3). Novel cases identified as individuals #30-#61.

**Supplemental table 2 – Behavioral and cognitive profile:** Behavioral phenotypical aspects of all known individuals with HAFOUS, also containing information about cognition (sorted by mutation type).

**Supplemental table 3 – Motor + speech development:** Milestones in motor and speech development for HAFOUS patients, categorized by mutation type.

**Supplemental table 4 – Musculoskeletal phenotype:** Musculoskeletal phenotypic features observed in HAFOUS patients, categorized by mutation type.

**Supplemental table 5 – Neurological phenotype**: Neurological manifestations in HAFOUS patients, sorted by mutation type.

**Supplemental table 6 – Endocrine phenotype:** Detailed endocrine phenotype of HAFOUS patients, sorted by mutation type.

**Supplemental table 7 – Respiratory phenotype:** Respiratory phenotype in HAFOUS patients, categorized by mutation type.

**Supplemental table 8 – Sleep + seizures + EEG**: Sleep patterns, EEG study results, and seizure occurrences in HAFOUS patients, sorted by mutation type.

**Supplemental table 9 – Eating behavior:** Parental-reported eating behavior of all known individuals with HAFOUS, sorted by mutation type.

**Supplemental table 10 – Gastrointestinal phenotype:** Gastrointestinal phenotypes in HAFOUS patients, categorized by mutation type.

**Supplemental table 11 – Body measurements:** Comprehensive body measurements of HAFOUS patients, sorted by mutation type.

**Supplemental table 12 – Birth and neonatal period:** Phenotypic aspects of the birth and neonatal period in HAFOUS patients, categorized by mutation type.

**Supplemental table 13 – More phenotypical aspects:** Further phenotypical aspects of HAFOUS patients, sorted by type of mutation. Also containing parental-reported symptoms not being listed in our questionnaire and parental-identified aspects being “most disabling” and “the most striking capability”.

**Supplemental table 14 – Severity score:** Calculation of severity scores for HAFOUS patients meeting missense or truncating mutation criteria within our cohort. Includes points per category, cumulative points, maximum achievable points, and severity scores per individual.

## Notes

### Competing Interest Statement

The authors have declared no competing interest.

### Funding Statement

This study did not receive any funding.

### Summary of Updates

We have received a variety of helpful comments from the reviewers and extensively adjusted our manuscript. In addition to a new figure, we have also included a correlation analysis of age and body measurements. Furthermore, we have re-evaluated the ACMG classification and made editorial changes to the manuscript.

