## Supplemental material for "Hao-Fountain Syndrome: 32 novel patients reveal new insights into the clinical spectrum"

**Supplemental tables**

🡪 excel file

**Supplemental methods**

*Calculation of the severity score*

Each participant's severity score needs to be calculated individually based on the answers provided in the questionnaire. In 18 categories, either one or no point was assigned, depending on the occurrence of symptoms. In three other categories, no, one, or two points could be achieved based on the severity of manifestation.

| Symptom | 0 | 1 | 2 |
| --- | --- | --- | --- |
| ASD | not prevalent | prevalent | not assigned |
| ADHD | not prevalent | prevalent | not assigned |
| ID | not prevalent | prevalent (IQ < 70) | not assigned |
| Motor development delay 🡪 milestones: free sitting, crawling, free walking | 1 or fewer motor milestone <90^th^ percentile^1^ | 2 or more motor milestones >90^th^ percentile^1^ | 2 or more motor milestones >99^th^ percentile^1^ |
| Speech development delay  🡪 milestones: first word, first combining words | Delay of no milestone >6 months^2^ | Delay of at least one milestone >6 months^2^ | Delay of both milestones >12 months^2^ |
| Hypotonia | not prevalent | prevalent | not assigned |
| Contractures | not prevalent | prevalent | not assigned |
| Motor dysfunction | No motor dysfunction | Motor dysfunction for 1/3 categories^3^ | Motor dysfunction for 3/3 categories |
| Speech dysfunction | not prevalent | prevalent | not assigned |
| Eating complications | not prevalent | prevalent (either hyperphagia or gaining weight difficulties) | not assigned |
| Chronic GI issues | not prevalent | prevalent (chronic constipation or diarrhea) | not assigned |
| Endocrine dysfunction | not prevalent | prevalent (GH deficiency or hypogonadism) | not assigned |
| Seizures | not prevalent | prevalent | not assigned |
| Sleep apnea | not prevalent | prevalent | not assigned |
| Eye anomalies | not prevalent | prevalent (any of the queried symptoms^4^) | not assigned |
| Abnormal threshold for pain | not prevalent | prevalent (high or low) | not assigned |
| Abnormal brain MRI | not prevalent | prevalent | not assigned |
| Short stature | not prevalent | prevalent (<3^rd^ percentile) | not assigned |
| Obesity | not prevalent | prevalent | not assigned |
| LGA or SGA | not prevalent | prevalent (<10^th^ or >90^th^ percentile for gestational age) | not assigned |
| Congenital anomalies | not prevalent | prevalent | not assigned |

According to WHO data

^2^ According to Dosman et al. 2012

^3^ We asked for gross, fine, and oral motor function

^4^ Farsightedness, nearsightedness, strabismus, cortical visual impairment, nystagmus, anisocoria

Finally, severity score was calculated by dividing achieved points by the maximum possible points to subtract unspecified categories from the calculation. This leads to a score ranging from 0 to 1, expressing the severest phenotype with a score of 1.

**Supplemental figures**

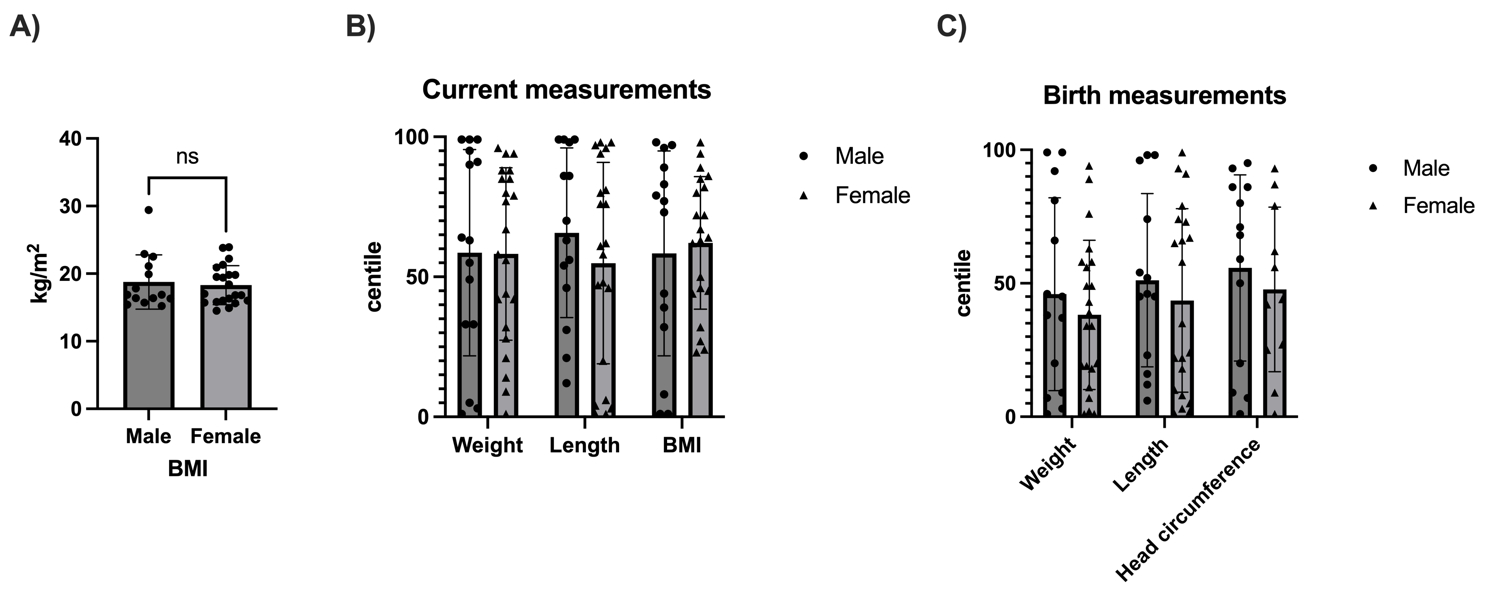

**Supplemental figure 1:** Body measurements compared by sex.

A: BMI values (Student's t-test); B: Current body measurement centiles; C: Birth body measurement centiles

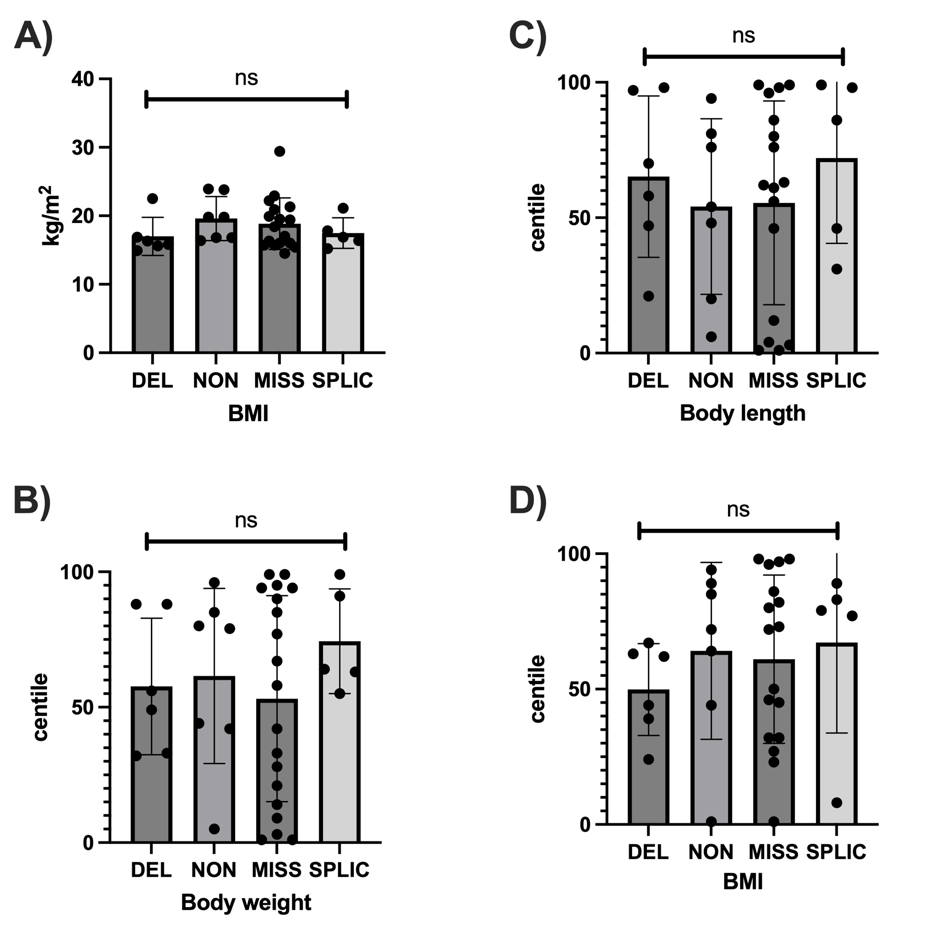

**Supplemental figure 2:** Current body measurements compared by genotype

A: BMI values (One-way ANOVA); B: Current body length centile (One-way ANOVA); C: Current body weight centile (One-way ANOVA); D: BMI centile (One-way ANOVA)

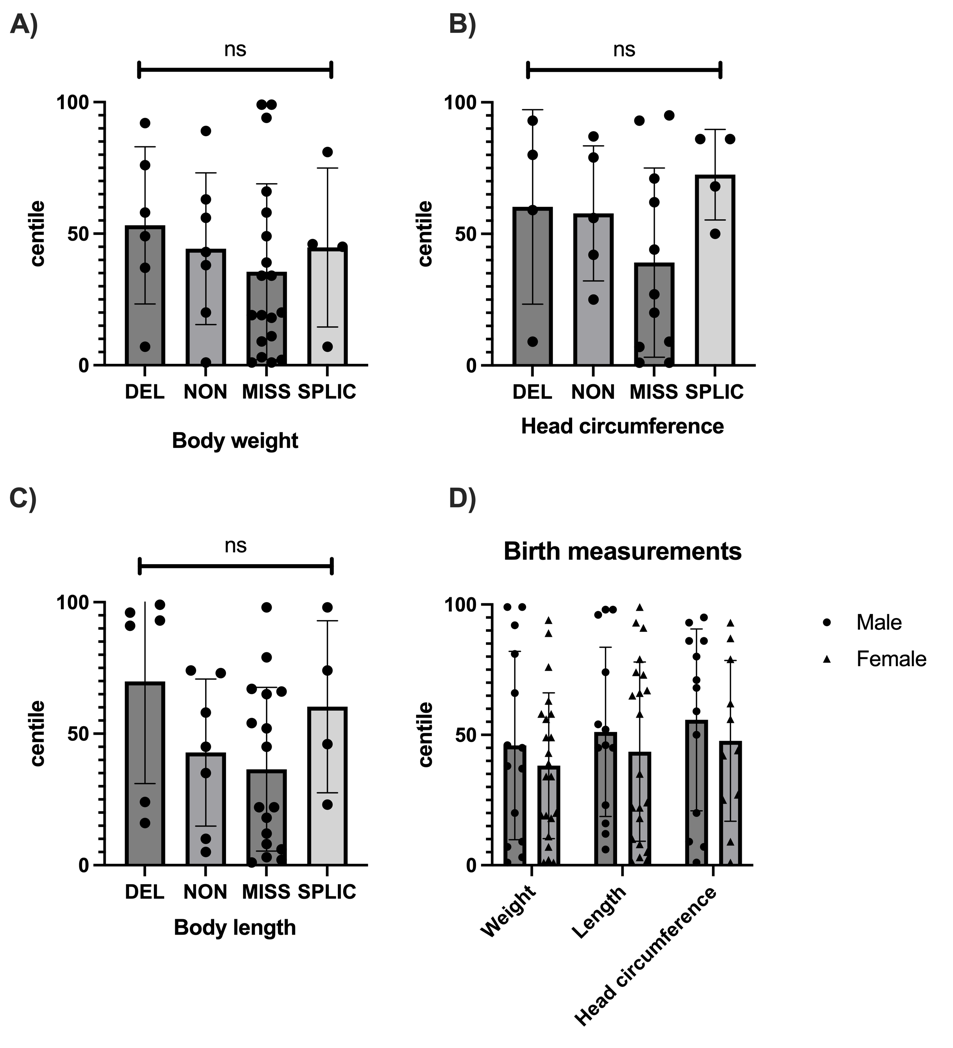

**Supplemental figure 3:** Birth body measurements compared by sex and genotype

A: Birth body weight centiles (One-way ANOVA); B: Birth head circumference centiles (One-way ANOVA); C: Birth body length centiles (One-way ANOVA); D: Birth measurements compared by sex

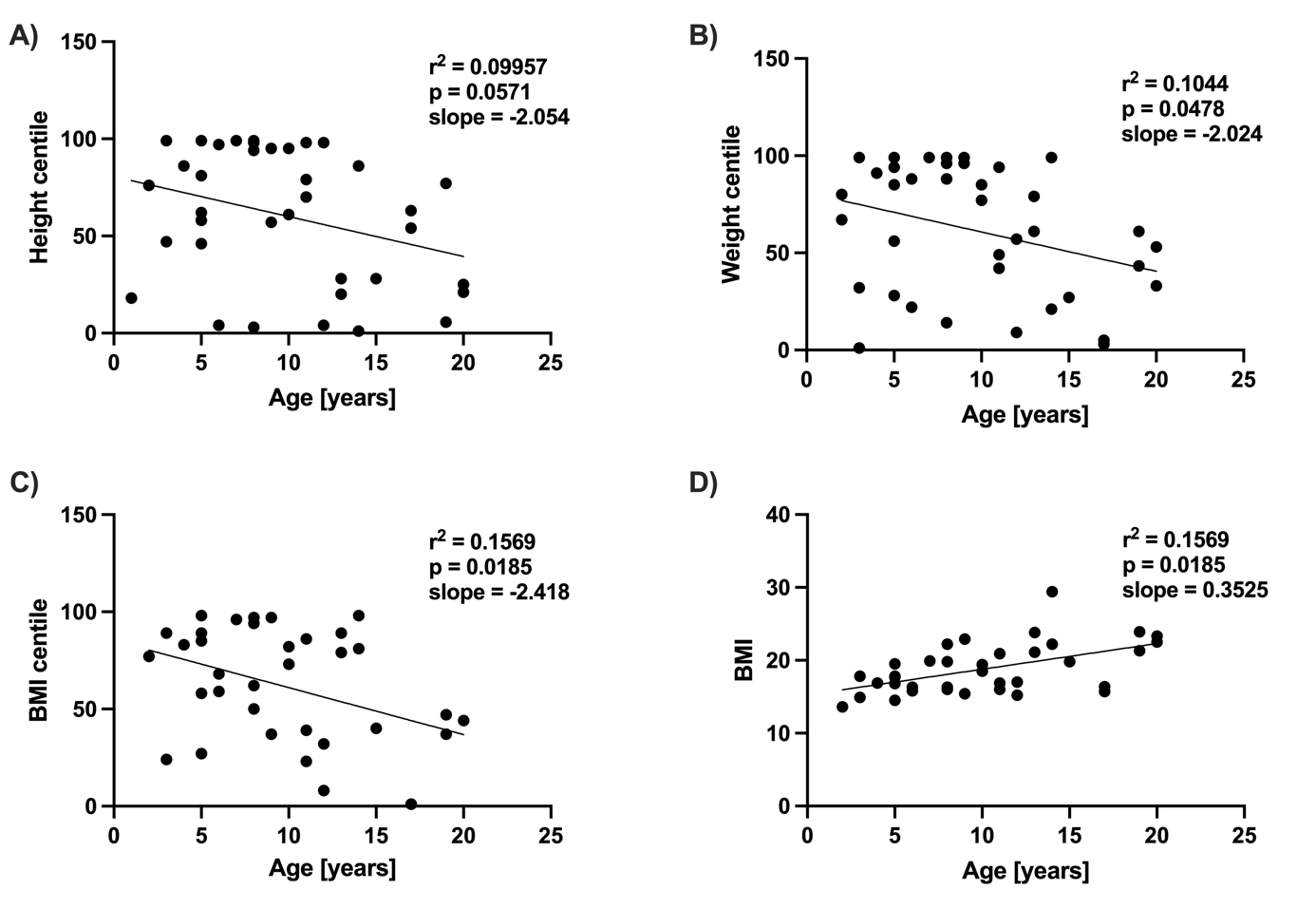

**Supplemental figure 4:** Body measurements plotted against age indicating a significant impact of age on weight centile (B), BMI centile (C), and BMI values (C)

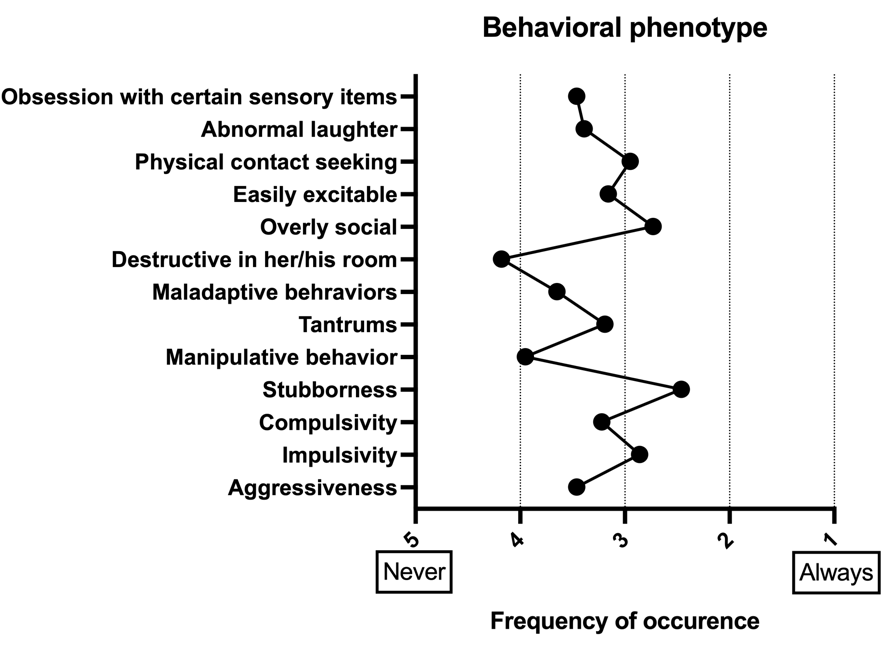

**Supplemental figure 5**: Behavioral traits of Hao-Fountain syndrome patients

Several behavioral traits and parental-reported frequency of occurrence ranging from 5 (Always) to 1 (Never) (n: 36).

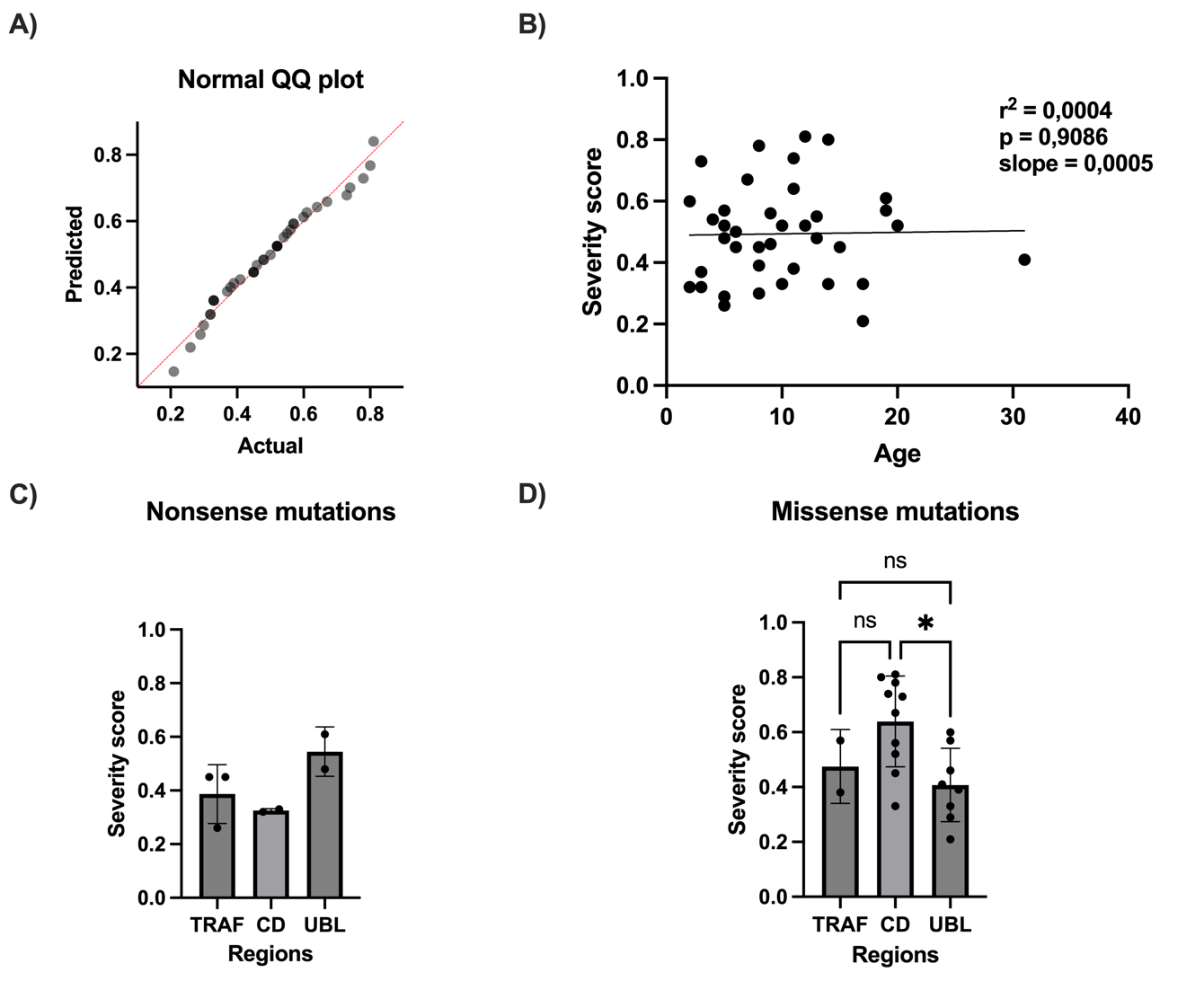

**Supplemental Figure 6:** Severity score QQ plot and assessment for different subgroups

A: QQ plot proofing normal distribution of severity scores within our cohort; B: Severity score plotted against age of individuals indicating no impact of age on severity; C: Severity score of patients with a truncating mutation plotted for the location of the predicted location of the truncation; D: Severity score of patients with a missense mutation plotted for the location of the predicted location of the truncation (Tukey's multiple comparison test following one-way ANOVA; CD vs. UBL: adjusted p-value = 0.0131)

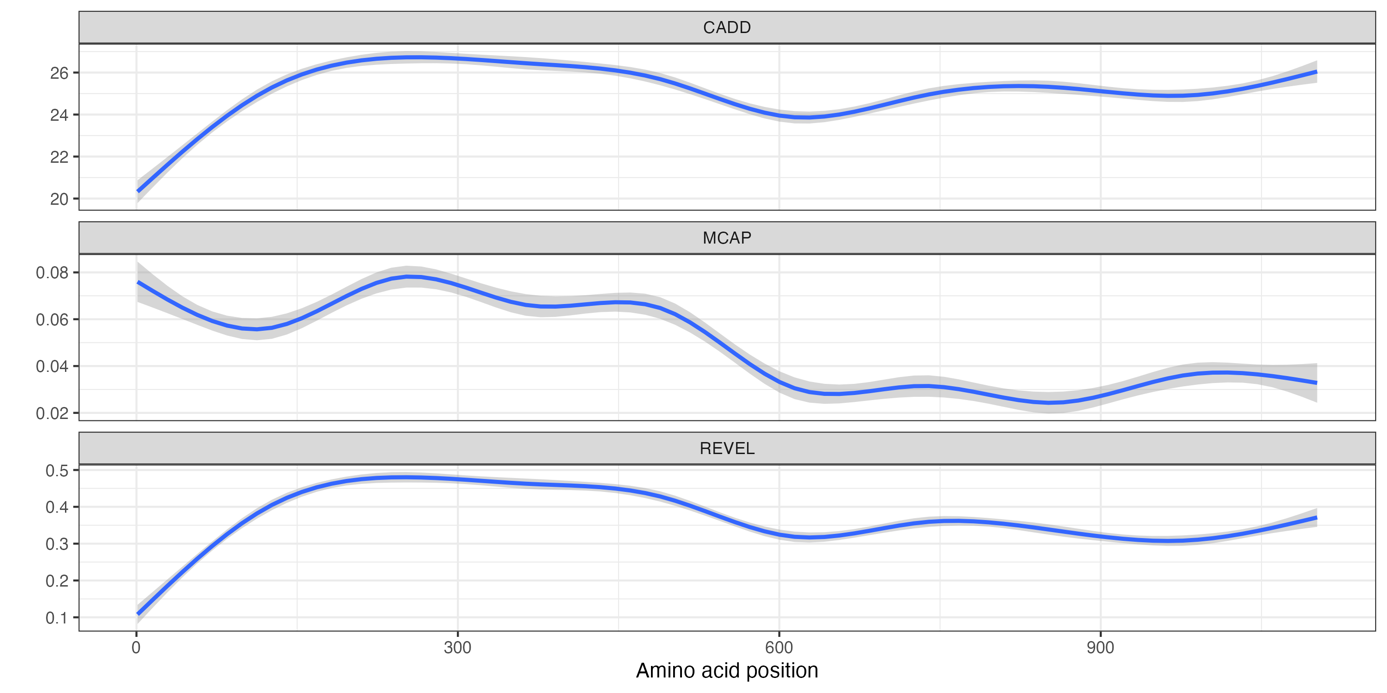

**Supplemental Figure 7**: Comparison of three pathogenicity score predictions

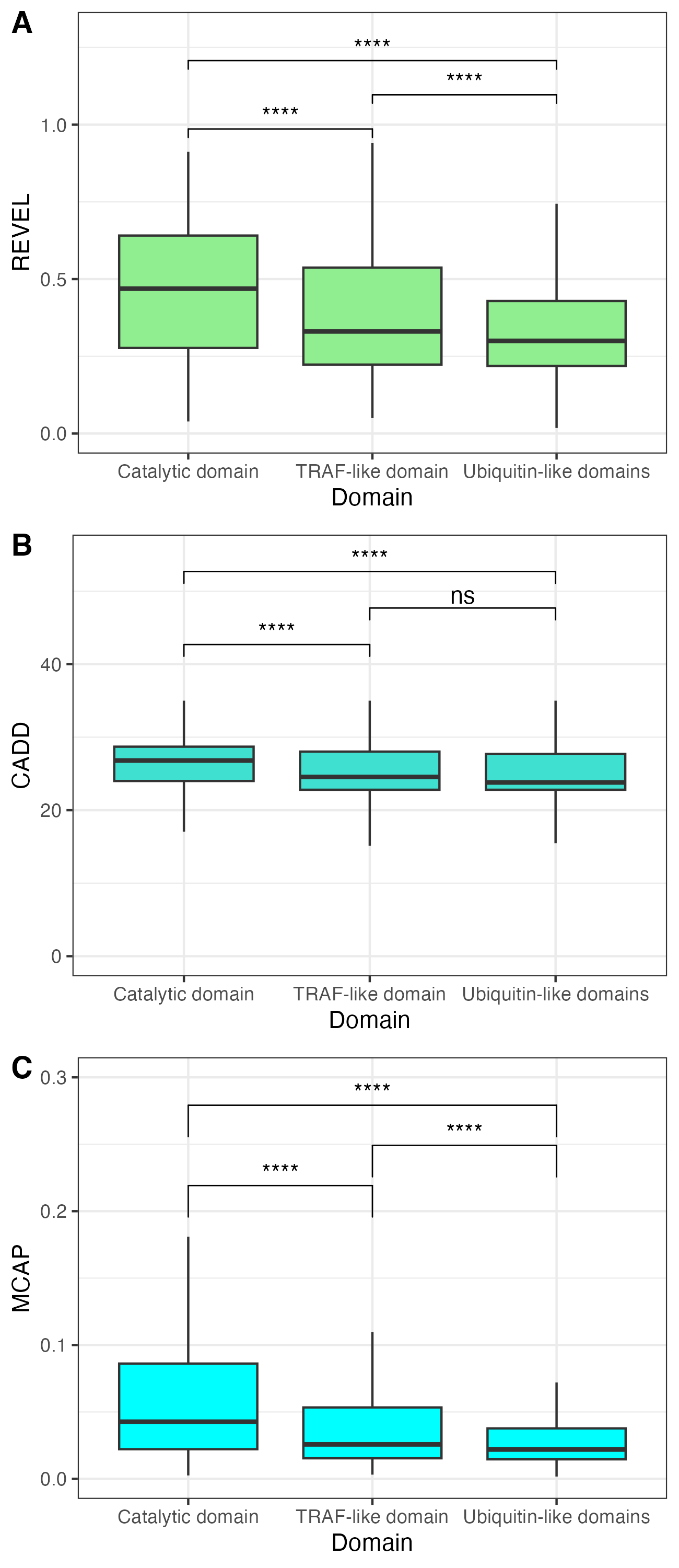

**Supplemental Figure 8:** Statistical comparison of domain-associated pathogenicity scores
