## Supplementary material for "Hao-Fountain Syndrome: 32 novel patients reveal new insights into the clinical spectrum": Questionnaire

Clinical questionnaire: **Hao-Fountain Syndrome** (USP7-related disorder)

*The following questionnaire is directed to parents of patients with the Hao-Fountain Syndrome.*
*If possible, patients are obviously very welcome to fill out this questionnaire on their own.*

**Personal information**

Current date (d/m/y): ____________________________________________________________

Your child's name: _______________________________________________________________

Your child's date of birth (d/m/y): ___________________________________________________

Current height (cm): ____________________ Date of measurement (d/m/y): __________________

Current weight (kg): ____________________ Date of measurement (d/m/y): __________________

*Regarding measurements, feel free to use other units (like lbs. and oz., ft. and in.), but please indicate which unit you used.*

Do you have a copy of the genetic testing report of your child? If so, could you share it with us?

- ☐ Yes, we have attached the genetic results.
- ☐ Yes, they have already been shared with Prof. Schaaf.
- ☐ No, we do not have genetic results.

How old was your child when the diagnosis Hao-Fountain-Syndrome was finally made? ___________

**Pregnancy/ Delivery/ Neonatal Period**

Gestational Age (at birth, weeks): ____________________________________

Maternal Age (at birth, weeks): _______________________________________

Paternal Age (at birth, weeks): _______________________________________

Type of delivery: ☐ Caesarean section ☐ vaginal delivery

Complications during pregnancy
- ☐ yes
  - ☐ polyhydramnios
  - ☐ subchorionic hemorrhage
  - ☐ others, please specify: ____________________________
- ☐ no
- ☐ unknown

Complications after delivery
- ☐ yes
  - ☐ difficulties maintaining temperature during birth
  - ☐ others, please specify: ____________________________
- ☐ no
- ☐ unknown

Birth measurements:

Length (cm): ________________________________________________________________

Birth weight (g): ____________________________________________________________

Head circumference (cm): ____________________________________________________

*Regarding measurements, feel free to use other units (like lbs. and oz., ft. and in.), but please indicate which unit you used.*

| | | | |
|---|---|---|---|
| Newborn hearing test: | ☐ passed | ☐ failed | ☐ unknown |
| Neonatal hypotonia (low muscle tone): | ☐ Yes | ☐ No | ☐ unknown |
| Neonatal poor suck: | ☐ Yes | ☐ No | ☐ unknown |
| Hydrocephalus as an infant: | ☐ Yes | ☐ No | ☐ unknown |
| Did you recognize decreased fetal movement? | ☐ Yes | ☐ No | ☐ unknown |

Congenital anomalies (e.g. heart defects, skeletal anomalies, **absent kidney**, **extreme overbite**; if possible please specify):

________________________________________________________________________________

________________________________________________________________________________

Please measure the **length of the hand and feet** of your child and record the results here.

Length of the foot: ___________________________________________________________________

Length of the hand, Part A: ____________________________________________________________

Length of the hand, Part B: ____________________________________________________________

*For reliable measuring of the foot please measure from the heel to the end of the big toe. The process of measuring the hand is illustrated in the graphic below.*

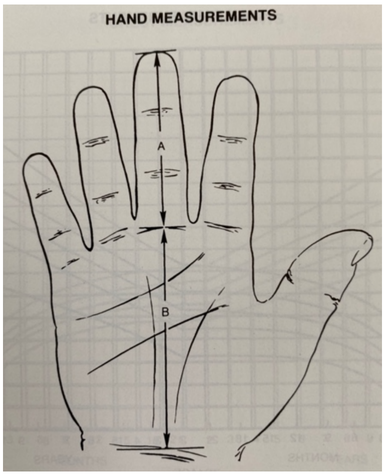


**<u>Developmental milestones (motor):</u>**

When was your child first able to **sit** independently (months)? ___________________________

When was your child first able to **crawl** (months)? ______________________________________

When was your child first able to **walk** independently (months)? __________________________

**<u>Developmental milestones (language):</u>**

When did your child speak the **first word** (other than “mama” or “dada”) (months)?

______________________

When was your child able to speak first **combining words** (months)?  _____________________

☐ So far, my child is nonverbal.

### Clinical features: Infant period/ Adult Stage

*For each of the following conditions, please tick yes if they apply to your child in the question. How have these clinical features evolved over time? Please note which symptom has improved/ got worse.*

#### 1. Cognitive evaluation

Needs special education? ☐ Yes ☐ No ☐ Unknown

Attends mainstream school but needs resources? ☐ Yes ☐ No ☐ Unknown

Formal cognitive evaluation (IQ or DQs)? ☐ Yes ☐ No ☐ Unknown

If your child did undergo any formal cognitive evaluation, are you willing to share the results with us?

O Yes, the results are attached

O I don't have them.

Judgement impaired? ☐ Yes ☐ No ☐ Unknown

(i.e. danger awareness limited)

#### 2. Muscular system and motor function

Hypotonia (beyond the neonatal period)?

☐ Yes ☐ No ☐ Unknown

Has the hypotonia evolved over time?

☐ Worsened

☐ Improved

☐ No change

☐ Additional comments:

Do you recognize an abnormal gait? ☐ Yes ☐ No ☐ unknown

Balance unsteadiness? ☐ Yes ☐ No ☐ unknown

Delayed/ impaired fine motor skills? ☐ Yes ☐ No ☐ unknown

Delayed/ impaired gross motor skills? ☐ Yes ☐ No ☐ unknown

Delayed/ impaired oral motor skills? ☐ Yes ☐ No ☐ unknown

(i.e. tongue thrusting, sucking/ swallowing disorders, frequent drooling, excessive chewing, lack of strength in the mouth)

Very flexible? (i.e. hyperextensible joints) ☐ Yes ☐ No ☐ unknown

**3. Behavioral features**

| | Always | | | | Never |
|---|---|---|---|---|---|
| Aggressiveness | ☐ | ☐ | ☐ | ☐ | ☐ |
| Impulsivity | ☐ | ☐ | ☐ | ☐ | ☐ |
| Compulsivity | ☐ | ☐ | ☐ | ☐ | ☐ |
| Stubbornness | ☐ | ☐ | ☐ | ☐ | ☐ |
| Manipulative behavior | ☐ | ☐ | ☐ | ☐ | ☐ |
| Lots of tantrums | ☐ | ☐ | ☐ | ☐ | ☐ |
| Maladaptive behaviors: biting, hair-pulling, pinching, slapping, yelling | ☐ | ☐ | ☐ | ☐ | ☐ |
| Destructive in her/ his room: empties drawers, tears books | ☐ | ☐ | ☐ | ☐ | ☐ |
| Overly social/ friendly | ☐ | ☐ | ☐ | ☐ | ☐ |
| Easily excitable | ☐ | ☐ | ☐ | ☐ | ☐ |
| Physical contact seeking | ☐ | ☐ | ☐ | ☐ | ☐ |
| Laughter abnormal/ inappropriate | ☐ | ☐ | ☐ | ☐ | ☐ |
| Obsession/ fascination with certain sensory items (i.e. water, crinkly items, mirrors, reflections) | ☐ | ☐ | ☐ | ☐ | ☐ |

**4. Eye anomalies**

Farsightedness? ☐ Yes ☐ No ☐ unknown

Nearsightedness? ☐ Yes ☐ No ☐ unknown

Strabismus or Esotropia (“crossed eyes”)? ☐ Yes ☐ No ☐ unknown

| | | | |
|---|---|---|---|
| Nystagmus (trembling of the eye)? | ☐ Yes | ☐ No | ☐ unknown |
| Cortical visual impairment? | ☐ Yes | ☐ No | ☐ unknown |
| Anisocoria (unequal size of the pupils)? | ☐ Yes | ☐ No | ☐ unknown |

**5. ASD**

| | | | |
|---|---|---|---|
| Any formal testing (ADOS, ADI-R)? | ☐ Yes | ☐ No | ☐ Unknown |
| Meets diagnostic criteria of autism in formal testing? | ☐ Yes | ☐ No | ☐ Unknown |
| Autistic features (without formal testing)? | ☐ Yes | ☐ No | ☐ Unknown |
| Obsessive compulsive behaviors? | ☐ Yes | ☐ No | ☐ Unknown |

Have the autistic features evolved over time?

☐ Worsened, _______________ got worse.

☐ Improved, _______________ has improved.

☐ No change

☐ Additional comments:

**6. ADHD**

| | | | |
|---|---|---|---|
| Attention deficit? | ☐ Yes, first diagnosed at age_____ | ☐ No | ☐ Unknown |
| Hyperactivity? | ☐ Yes, first diagnosed at age_____ | ☐ No | ☐ Unknown |
| Formal diagnosis of ADHD? | ☐ Yes, first diagnosed at age_____ | ☐ No | ☐ Unknown |
| Behavioral problems? | ☐ Yes, first diagnosed at age_____ | ☐ No | ☐ Unknown |

If you ticked yes, have they evolved? Please note which one has improved/ got worse.

☐ Worsened, _______________ got worse.

☐ Improved, _______________ has improved.

☐ No change

Additional comments:

**7. Diagnostic studies**

| | | | |
|---|---|---|---|
| Brain MRI? | ☐ Yes | ☐ No | ☐ Unknown |
| Thin/ hypoplastic corpus callosum on brain MRI? | ☐ Yes | ☐ No | ☐ Unknown |
| EEG? | ☐ Yes | ☐ No | ☐ Unknown |

If you ticked yes, are you willing to share the results with us?

☐ Yes, the results are attached.

☐ I don't have them.

**8. Neurology**

| | | | |
|---|---|---|---|
| Low threshold for pain? | ☐ Yes | ☐ No | ☐ Unknown |
| High threshold for pain? | ☐ Yes | ☐ No | ☐ Unknown |
| Noticeable very ticklish (i.e. shower tickles)? | ☐ Yes | ☐ No | ☐ Unknown |
| Significant limitation in communication skills? | ☐ Yes | ☐ No | ☐ Unknown |
| Reduced expressive communication? | ☐ Yes | ☐ No | ☐ Unknown |
| Reduced receptive communication? | ☐ Yes | ☐ No | ☐ Unknown |
| Non-verbal communication through facial expression, body postures and gestures? | ☐ Yes | ☐ No | ☐ Unknown |

**9. Endocrine system**

| | | | |
|---|---|---|---|
| Hypothyroidism? | ☐ Yes | ☐ No | ☐ Unknown |
| Hyperthyroidism? | ☐ Yes | ☐ No | ☐ Unknown |
| Adrenal insufficiency? | ☐ Yes | ☐ No | ☐ Unknown |
| Growth hormone insufficiency? | ☐ Yes | ☐ No | ☐ Unknown |
| Panhypopituitarism ("Panhypopit")? | ☐ Yes | ☐ No | ☐ Unknown |

**10. Hypogonadism**

Was hypogonadism diagnosed? ☐ Yes ☐ No ☐ Unknown

a) For men/ boys:

cryptorchidism (undescended testicle) ☐ Yes ☐ No ☐ Unknown

micropenis ☐ Yes ☐ No ☐ Unknown

b) For women/ girls:

delayed development of secondary sex characteristics ☐ Yes ☐ No ☐ Unknown

**11. Respiratory system**

Required intubation during lifetime? ☐ Yes ☐ No ☐ Unknown

Required mechanical ventilation during lifetime? ☐ Yes ☐ No ☐ Unknown

Required a tracheostomy during lifetime? ☐ Yes ☐ No ☐ Unknown

Asthma? ☐ Yes ☐ No ☐ Unknown

**12. Sleep**

Sleep disturbances? ☐ Yes ☐ No ☐ Unknown
(i.e. problems initiating, maintaining sleep)

Sleep apnea? ☐ Yes ☐ No ☐ Unknown
(Pauses in breathing or periods of shallow breathing during sleep)

Needs much rest (i.e. more than siblings) ☐ Yes ☐ No ☐ Unknown

Tires easily/ rapid loss of energy ☐ Yes ☐ No ☐ Unknown

**13. Seizures**

Seizures? ☐ Yes ☐ No ☐ Unknown

How many?________________ / year

Medications? ____________________________

Infantile spasms? ☐ Yes, first appeared at age_____ ☐ No ☐ Unknown

Febrile seizures? ☐ Yes, first appeared at age_____ ☐ No ☐ Unknown

Diagnosed Panayiotopoulos Syndrome? ☐ Yes, first diagnosed at age_____ ☐ No ☐ Unknown
i.e. early onset childhood occipital epilepsy

If you ticked yes, have the seizures evolved?

☐ Worsened

☐ Improved

☐ No change

☐ Additional comments:

**14. Eating**

| | | | |
|---|---|---|---|
| Feeding difficulties in infancy? | ☐ Yes | ☐ No | ☐ Unknown |
| Eating difficulties in childhood and at adult age? | ☐ Yes | ☐ No | ☐ Unknown |
| Hyperphagia? | ☐ Yes | ☐ No | ☐ Unknown |

(i.e. excessive eating, food-seeking behavior, excessive weight gain)

Onset?
☐ Neonatal period (first month of life)
☐ Infancy (second month - two years)
☐ Early childhood (3 – 8 years)
☐ Other: ______________________

| | | | |
|---|---|---|---|
| Difficulty gaining weight? | ☐ Yes | ☐ No | ☐ Unknown |
| Mouthing behavior? | ☐ Yes | ☐ No | ☐ Unknown |
| Chewing inedible objects? | ☐ Yes | ☐ No | ☐ Unknown |
| Remarkable appetite for dairy products? | ☐ Yes | ☐ No | ☐ Unknown |

**15. GERD: Gastroesophageal reflux disease**

Was GERD diagnosed? ☐ Yes ☐ No ☐ Unknown

Please tick the frequency of the following symptoms:

| | *daily* | *weekly* | *monthly* | *all 3 months* | *very rare/ never* |
|---|---|---|---|---|---|
| Heartburn | ☐ | ☐ | ☐ | ☐ | ☐ |
| Regurgitation of food or sour liquid | ☐ | ☐ | ☐ | ☐ | ☐ |
| Nasea/ vomiting | ☐ | ☐ | ☐ | ☐ | ☐ |

**16. Skeletal system**

Scoliosis or kyphosis? ☐ Yes ☐ No ☐ Unknown

Joint contractures? ☐ Yes ☐ No ☐ Unknown

If you ticked yes, where? ______________________________________________________

Hip dysplasia? ☐ Yes ☐ No ☐ Unknown

Dental issues? ☐ Yes ☐ No ☐ Unknown

(i.e. abnormal dentition, large front teeth)

If you ticked yes, please describe these issues: ___________________________________________

**17. Gastrointestinal symptoms**

Chronic constipation/ chronic diarrhea? ☐ Yes ☐ No ☐ Unknown

Incontinence inappropriate for her/ his age? ☐ Yes ☐ No ☐ Unknown

**18. Other features**

Does your child pick at her/his skin? ☐ Yes ☐ No ☐ Unknown

Hearing difficulties? ☐ Yes ☐ No ☐ Unknown

Temperature instability? ☐ Yes ☐ No ☐ Unknown
(e.g. abnormal heat sensitivity)

Hypercalcemia? ☐ Yes ☐ No ☐ Unknown

Jaundice? ☐ Yes ☐ No ☐ Unknown

Polycystic kidney disease? ☐ Yes ☐ No ☐ Unknown

Skin remains pale despite sun exposure? ☐ Yes ☐ No ☐ Unknown

Frequent eczema? ☐ Yes ☐ No ☐ Unknown

Did you observe any other features?

______________________________________________________________________________

______________________________________________________________________________

______________________________________________________________________________

From your perspective, what is the most disabling symptom that your daughter/ son presents that is specifically related to her/ his disease?

______________________________________________________________________________

From your perspective, what is the most striking capability that your daughter/ son presents that is specifically related to her/ his disease?

______________________________________________________________________________

Are you willing to share pictures with us of the different stages of life?

☐ Yes, pictures are attached. ☐ No, I don’t want to share them.

Do you agree that photographs and medical information of your child may be used for automated analysis with “Face2Gene”, without sharing your child’s name? (www.face2gene.com)

☐ Yes, the signed consent form is attached. ☐ No.
